# Knowledge and attitudes of caregivers of under-five children toward the malaria vaccine in the Tiko Health District, Cameroon: A community-based cross-sectional study

**DOI:** 10.1101/2025.04.30.25326760

**Authors:** Idang Maureen Abiache, Divine Nsobinenyui, Chrisantus Eweh Ukah, Yunika Larissa Kumenyuy, Ngu Claudia Ngeha, Randolf Wefuan, Syveline Zuh Dang, Ndip Esther Ndip, Mirabelle Pandong Feguem, Dickson S. Nsagha

## Abstract

**Background:** Despite progress in malaria control, it remains a significant public health burden in sub-Saharan Africa, especially among children under five. The introduction of the RTS,S/AS01 (Mosquirix) malaria vaccine offers hope; however, its successful implementation depends largely on caregivers’ knowledge and attitudes. This study assessed the knowledge and attitudes of caregivers of under-five children regarding the malaria vaccine in the Tiko Health District of Cameroon.

**Methods:** A descriptive cross-sectional survey was conducted from February to April 2025 in four randomly selected health areas within the Tiko Health District. A total of 410 caregivers of children aged 0–5 years were recruited using a multistage sampling technique. Data were collected using a structured, pre-tested questionnaire and analyzed using SPSS version 26. Descriptive statistics summarized sociodemographic characteristics, and knowledge and attitude scores were computed using a scoring system, with a 60% threshold used to define adequate knowledge and positive attitudes.

**Results:** The mean age of participants was 33.6 years (SD = 8.9), and the majority were female (83.2%). Although 60.7% of caregivers had heard about the malaria vaccine, only 26.6% demonstrated good knowledge, and 25.1% exhibited positive attitudes toward it. Healthcare providers were the most common source of information (35.4%). Misconceptions about vaccine efficacy and purpose were prevalent, and awareness of the vaccine’s approval in Cameroon was limited (41.8%). Positive attitudes were associated with trust in healthcare workers and belief in the vaccine’s importance for child health.

**Conclusion:** Caregivers in the Tiko Health District exhibited low levels of knowledge and generally negative attitudes toward the malaria vaccine. These findings highlight the need for targeted health education interventions and community engagement strategies to improve vaccine acceptance and uptake in this high-risk population.

**What is known on the topic:**

- Malaria remains a leading cause of morbidity and mortality among children under five in sub-Saharan Africa, despite the availability of preventive tools such as insecticide-treated nets and chemoprevention.
- The RTS,S/AS01 (Mosquirix) malaria vaccine has been approved by the WHO as an additional tool to reduce malaria incidence in young children in endemic areas.
- Successful vaccine uptake depends significantly on caregivers’ knowledge and attitudes, which influence their willingness to seek vaccination for their children.

**What this study adds:**

- This study reveals that although 60.7% of caregivers in the Tiko Health District had heard of the malaria vaccine, only 26.6% demonstrated good knowledge, and just 25.1% had positive attitudes toward it.
- Misconceptions regarding the vaccine’s purpose, effectiveness, and safety were prevalent, and over one-third of caregivers reported receiving no information at all on the vaccine.
- These findings underscore the urgent need for targeted, community-based health education and communication strategies to improve caregivers’ understanding and acceptance of the malaria vaccine in high-risk settings.

## Background

Malaria remains one of the most significant public health challenges globally, particularly in sub-Saharan Africa[1,2][3] where it disproportionately affects vulnerable populations, including children under five years of age[4]. According to the World Health Organization (WHO), malaria accounted for an estimated 619,000 deaths in 2021[5], with young children representing a substantial proportion of these fatalities. Despite ongoing efforts to control malaria through preventive measures such as insecticide-treated nets and antimalarial medications, the disease continues to pose a severe threat to child health in endemic regions[6,7].

The development of malaria vaccines has emerged as a promising strategy to complement existing prevention and treatment measures[8,8]. The RTS,S/AS01 (Mosquirix) vaccine, the first malaria vaccine to receive a WHO recommendation for use in children[9,10], has shown efficacy in reducing malaria incidence and severe disease in clinical trials[11,12]. However, the successful implementation of vaccination programs is contingent upon the knowledge and attitudes of caregivers, who play a crucial role in the health-seeking behaviors of their children[13–15].

In the Tiko health district, where malaria transmission is high[16], understanding caregivers’ perceptions and knowledge about the malaria vaccine is essential for developing effective public health strategies. Caregivers’ attitudes towards vaccination can significantly influence vaccine uptake and adherence[17,18], ultimately affecting the overall impact of vaccination campaigns. Factors such as cultural beliefs, previous experiences with healthcare services, and access to information can shape these attitudes and knowledge levels.

Research has indicated that misinformation and lack of awareness about vaccines can hinder vaccination efforts[19,20], leading to lower coverage rates. Moreover, caregivers’ socio-economic status, education level, and exposure to health education initiatives can further affect their understanding and acceptance of new interventions like the malaria vaccine. Therefore, assessing caregivers’ knowledge and attitudes towards the malaria vaccine in the Tiko Health District is critical for identifying barriers to vaccination and informing tailored educational interventions.

This study aims to fill the gap in knowledge regarding caregivers’ perspectives on the malaria vaccine for under-five children in the Tiko health district. By exploring these dimensions, we hope to contribute valuable insights that can enhance vaccination strategies and ultimately reduce the burden of malaria among young children in this high-risk area.

## Materials And Methods

### Research Design

This study used a descriptive cross-sectional survey design to assess the knowledge and attitudes of caregivers of children age 0-5 years in the Tiko Health District

### Study Area

The study was carried out in the Tiko Health District of the Southwest Region of Cameroon from 1^st^ of March 2025 to the 11^th^ of April 2025. Tiko Health District (THD), found in Fako Division, South West Region of Cameroon is located between latitude 9°32’2“N to 9°40’9”N and longitude 9°25’7“E to 9°55’7”E and an altitude of 32.84 m (107.76 ft). Tiko District, located in the Southwest Region of Cameroon, is divided into eight health areas. These health areas are managed by the local health administration and serve as the primary zones for delivering healthcare services to the community. Each health area typically includes a number of health centers and posts that provide essential services, such as maternal and childcare, immunization, treatment for infectious diseases like malaria, and other public health services.

### Study Settings and Duration

This study was conducted in some selected health areas of the Tiko Health District from January 2025 to March 2025.

### Target Population

The target population for this study was caregivers of children 0 to 5 years old resident within the Tiko Health District.

### Sample Size

The sample size was obtained using the formula for estimation of confidence interval for a proportion.

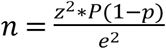

Where;

Z= 1.96 (for 95% confidence level)

P= 0.5 (estimated proportion),

e= 0.05(margin of error)

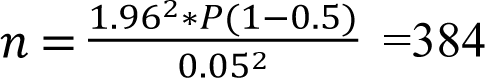. This was made up to 410 participants

### Sampling Technique

This study employed a multi-stage sampling technique to assess the knowledge and attitudes of caregivers of under-five children toward the malaria vaccine within the Tiko health districts. The Tiko health districts comprise eight distinct health areas. To ensure a representative sample while maintaining feasibility, four of these health areas were randomly selected using a simple random sampling method. This approach allowed for an unbiased selection process, ensuring that each health area had an equal opportunity to be included in the study. Following the selection of the health areas, community health workers were engaged to identify households with children under the age of five. These health workers possess intimate knowledge of their respective communities and were instrumental in locating eligible households. The identification process involved collaboration with community health workers who provided a list of households with under-five children within the selected health areas.

Once the households were identified, caregivers of under-five children were approached for participation in the study. The selection of participants was based on their availability and willingness to participate, ensuring that informed consent was obtained prior to data collection. This systematic approach facilitated the recruitment of a diverse sample of caregivers, thereby enhancing the reliability and validity of the findings regarding their knowledge and attitudes toward the malaria vaccine. The sampling technique utilized in this study involved a combination of random selection of health areas and purposeful identification of households by community health workers, which collectively contributed to a robust methodology for understanding caregiver perspectives in the Tiko health districts.

### Inclusion Criteria

Caregivers of under-five children living in the Tiko health district who gave their consent were included in the study.

### Exclusion Criteria

Under-five children’s caregivers who were severely ill at the time of data collection.

## Data Collection Tools and Methods

Data collection for this study was conducted using a pre-tested structured questionnaire designed to gather comprehensive information on the knowledge and attitudes of caregivers of under-five children toward the malaria vaccine. The questionnaire was systematically divided into four distinct sections to facilitate focused data collection:

## Section A: Socio-Demographic Variables

This section collected essential demographic information, including age, level of education, religion, location, and marital status of the caregivers. These variables provide context for understanding the background of participants and their potential influence on knowledge and attitudes toward the malaria vaccine.

## Section B: Knowledge of Caregivers

This section assessed the knowledge of caregivers regarding the malaria vaccine through ten closed-ended questions. These questions were designed to evaluate caregivers’ understanding of malaria, its transmission, prevention strategies, and specific details about the malaria vaccine.

## Section C: Attitudes Toward the Malaria Vaccine

This section focused on the attitudes of caregivers toward the malaria vaccine, comprising ten closed-ended questions. The questions aimed to capture caregivers’ perceptions, beliefs, and feelings about the vaccine, including any concerns or misconceptions they may have.

To ensure the validity and reliability of the questionnaire, a pre-test was conducted among ten under-five children in the Buea Health District at the Tole Health Area. This pre-test allowed for adjustments to be made to improve clarity and applicability during data collection.

## Data Collection

Data collection was carried out by the principal investigator and four trained research assistants using the structured questionnaires. The process involved both self-administration for caregivers who were literate and interviewer administration for those who required assistance. For literate participants, the questionnaires were self-administered; for those who were unable to read or write, the research assistants read the questions aloud and recorded their responses.

Prior to participation, all caregivers were adequately informed about the study through a written information sheet and detailed verbal explanations. Written informed consent was obtained from each participant before proceeding with data collection. Participants were made aware of their rights to withdraw from the study at any time without any repercussions. Confidentiality was strictly maintained by anonymizing responses; no names were recorded on the questionnaires. Instead, each questionnaire was assigned a unique file number, accessible only to the investigator for data analysis purposes.

The research assistants underwent a comprehensive training program that included a detailed training agenda and manual. Training covered essential topics such as data collection techniques, community engagement strategies, ethical considerations, and maintaining confidentiality throughout the study process. This training ensured that all research assistants were well-prepared to conduct data collection effectively and ethically.

## Data Management and Data Analysis

Data management and analysis followed a systematic approach to ensure accuracy and reliability. Upon collection, questionnaires were thoroughly checked for completeness. Any incomplete questionnaires were discarded to maintain the integrity of the data.

The completed questionnaires were securely stored in a locked cupboard, accessible only to the principal investigator, until the data collection process was finalized. Once data collection was complete, an Excel spreadsheet generated from Kobo Toolbox was imported into SPSS version 26 for analysis. Additionally, a soft copy of the spreadsheet was saved on a flash drive and sent via email for backup purposes.

Data analysis was conducted using the Statistical Package for the Social Sciences (SPSS) software version 26, with results presented in tables and charts. Continuous variables, such as age, were described using summary statistics, including means and standard deviations. Categorical variables, such as educational level and marital status, were summarized using frequency tables and pie charts.

To assess caregivers’ knowledge and attitudes, toward the malaria vaccine, a scoring system was implemented. Each of the knowledge and attitudes sections of the questionnaire contained ten questions, with a maximum obtainable score of 10/10. Correct answers received a score of one (1), while incorrect answers received zero (0). The total score for each participant was calculated based on their responses. The average score across all participants for each section was determined; those scoring at or above the average were classified as having good knowledge, attitudes, or practices, while those scoring below the average were classified as having poor knowledge, attitudes, or practices.

## ETHICAL CONSIDERATIONS

Ethical clearance was obtained from the Ethics Committee for Human Health Research in the Southwest Region of Buea. Additionally, administrative authorizations were secured from the Faculty of Health Sciences Institutional Review Board at the University of Douala, which were subsequently submitted to the Tiko District Health Services prior to data collection.

Informed consent was obtained from all participants prior to their inclusion in the study. Participants were provided with a detailed consent form that outlined the purpose of the research, the procedures involved, potential risks and benefits, and their right to withdraw from the study at any time without any consequences. This ensured that participants had a clear understanding of their involvement and could make an informed decision regarding their participation.

Confidentiality was strictly maintained throughout the research process. All data collected were anonymized, and identifying information was removed to protect participants’ privacy. Data were stored securely and accessible only to authorized research personnel. Findings were reported in aggregate form, ensuring that individual responses could not be traced back to any participant. These measures were implemented to foster trust and ensure that participants felt safe and secure while contributing to the research.

## RESULTS SOCIO-DEMOGRAPHIC CHARACTERISTICS

The mean age of the 410 caregivers was 33.6 and the standard deviation was 8.9. A total of 219 (53.4%) of caregivers were within the age group 21-35 years and 341 (83.2%) of them were females. Secondary education was the dominant educational level 160 (39.0%) and 340 (82.9%) were Christians. A majority 249 (60.7%) were married and 158 (38.5%) were self-employed. A vast majority 329 (80.2%) of the caregivers were direct parents of the under children and 391 (95.4%) were non-smokers (Table 1)

**Table 1:**
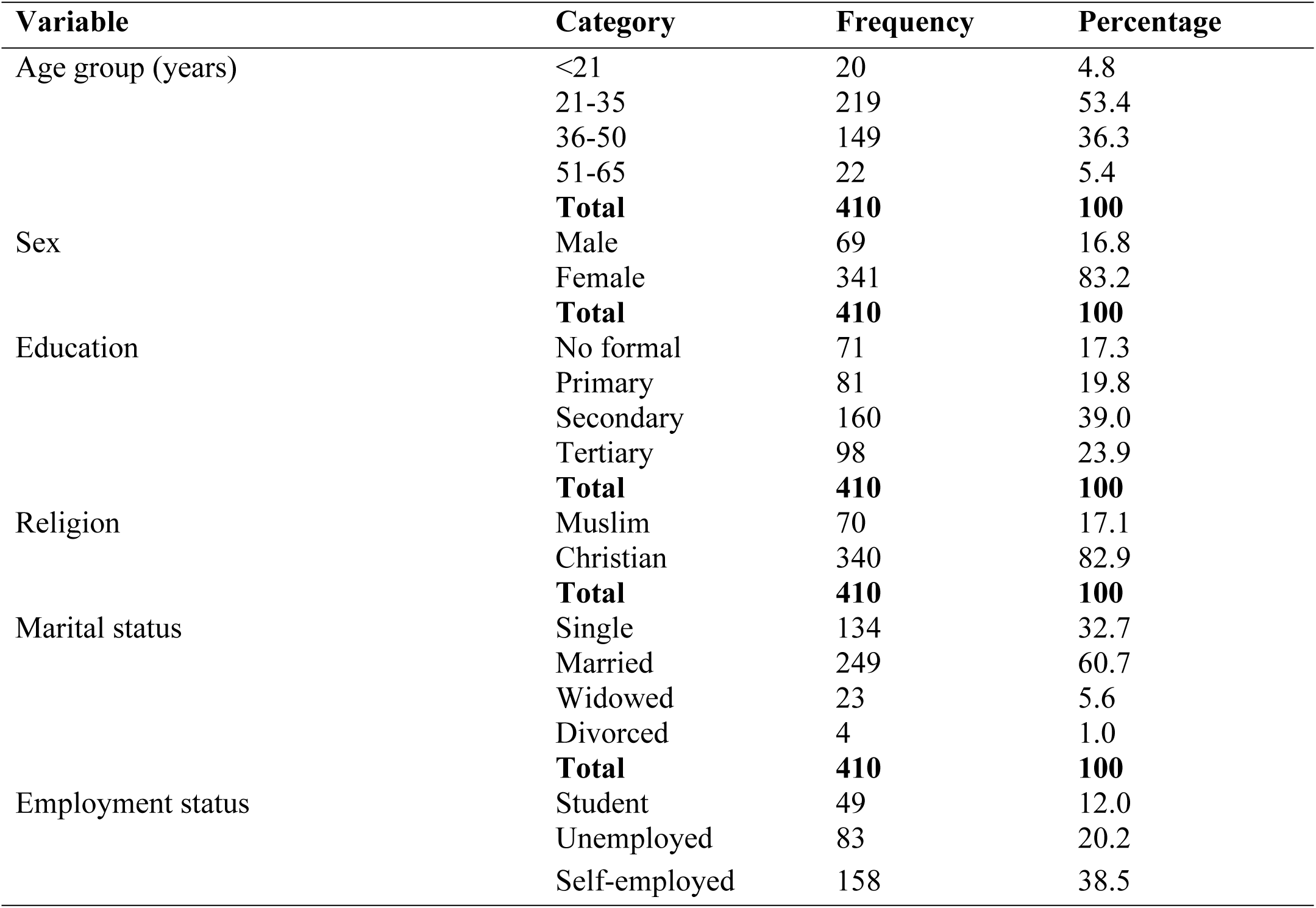
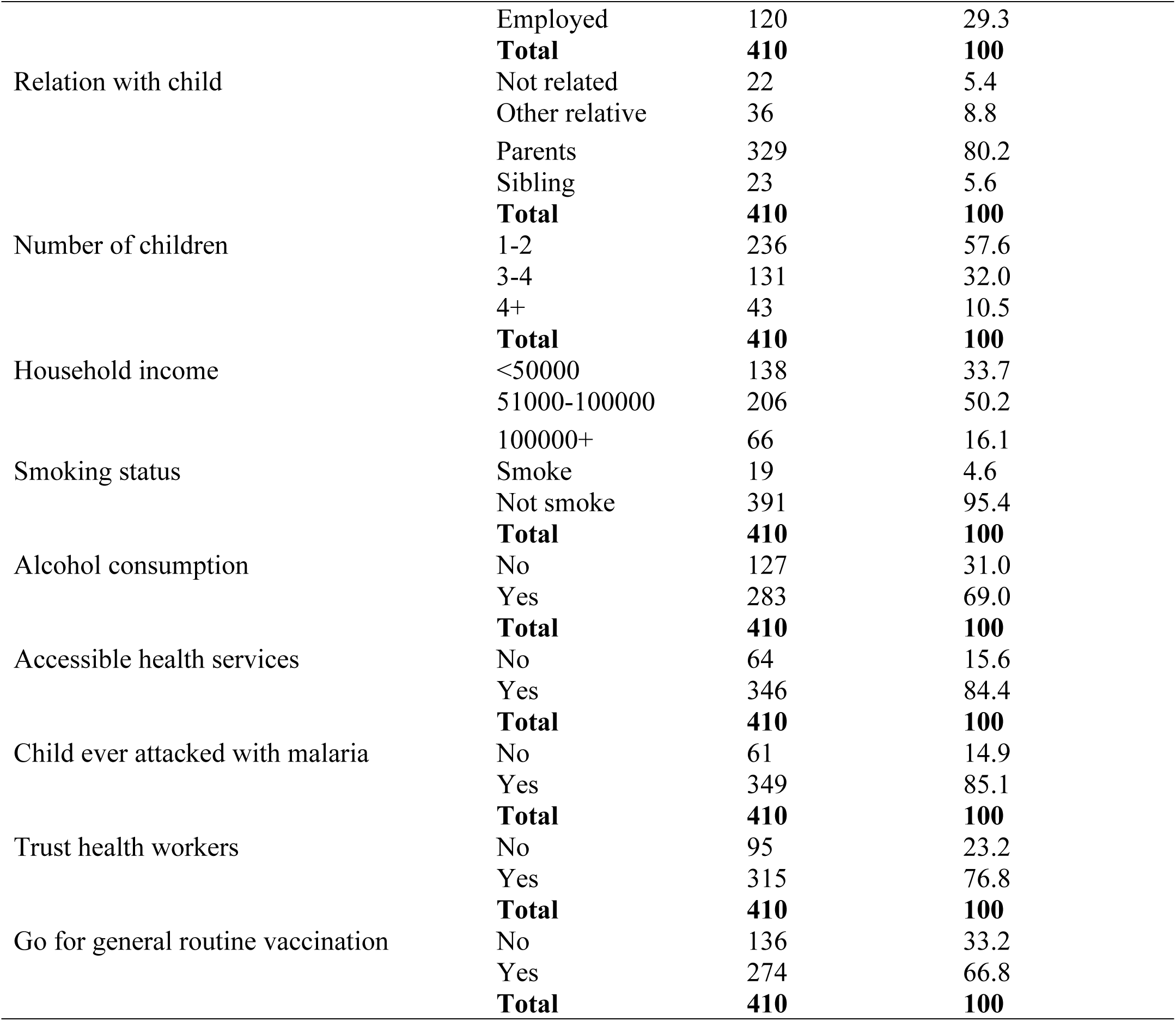
Socio-demographic characteristics of caregivers

### Knowledge of Caregivers of Under Five Children on the Malaria Vaccine in the Tiko Health District

Regarding the knowledge of caregivers on the malaria vaccine (Table 2), 145 (35.4%) sources of information on the vaccine was healthcare worker and 249 (60.7%) were aware of the existence of the malaria vaccine. Of the 249 who were aware of the existence of the malaria vaccine, 145 (58.2%) did not know that the vaccine was approved in Cameroon and 109 (43.8%) knew that the purpose of the malaria vaccine was to boost immunity in order to prevent the malaria infection.

**Table 2:**
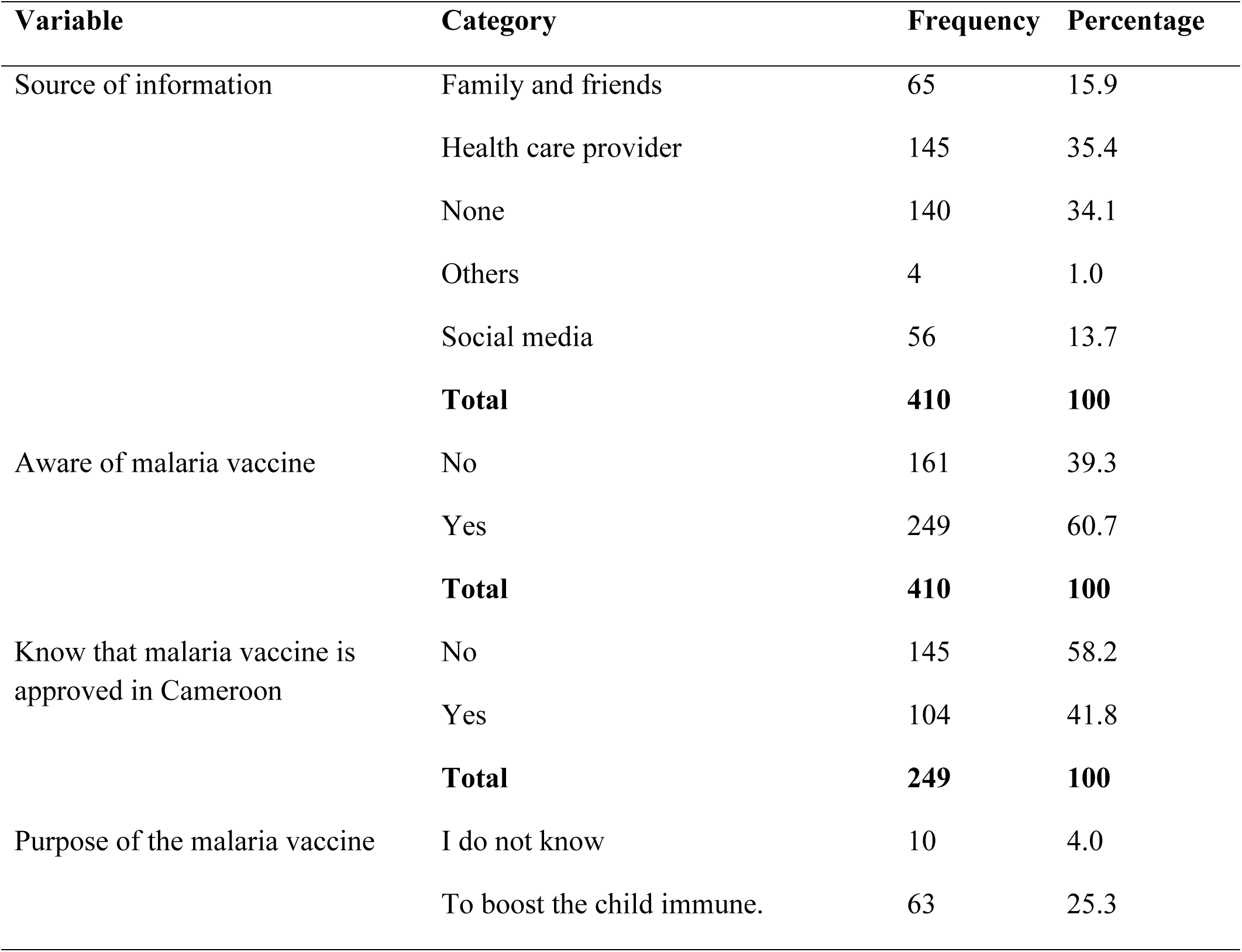
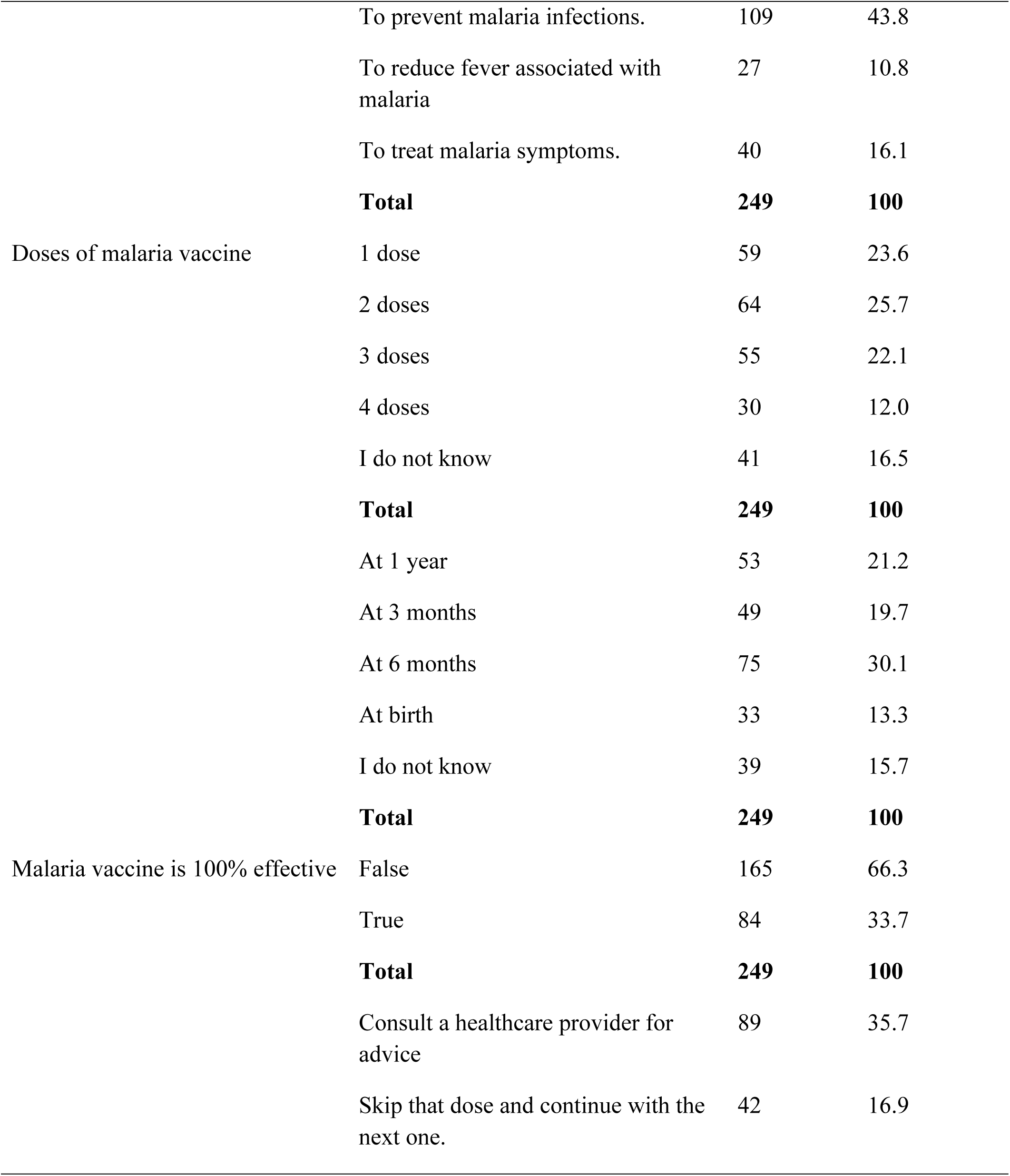
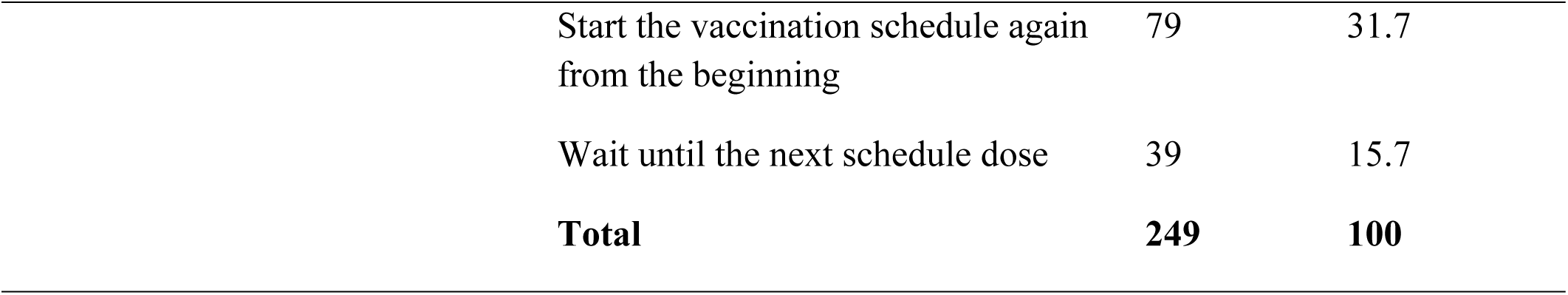
Knowledge of under-five caregivers on the malaria vaccine in the Tiko Health District

**Figure 1:**
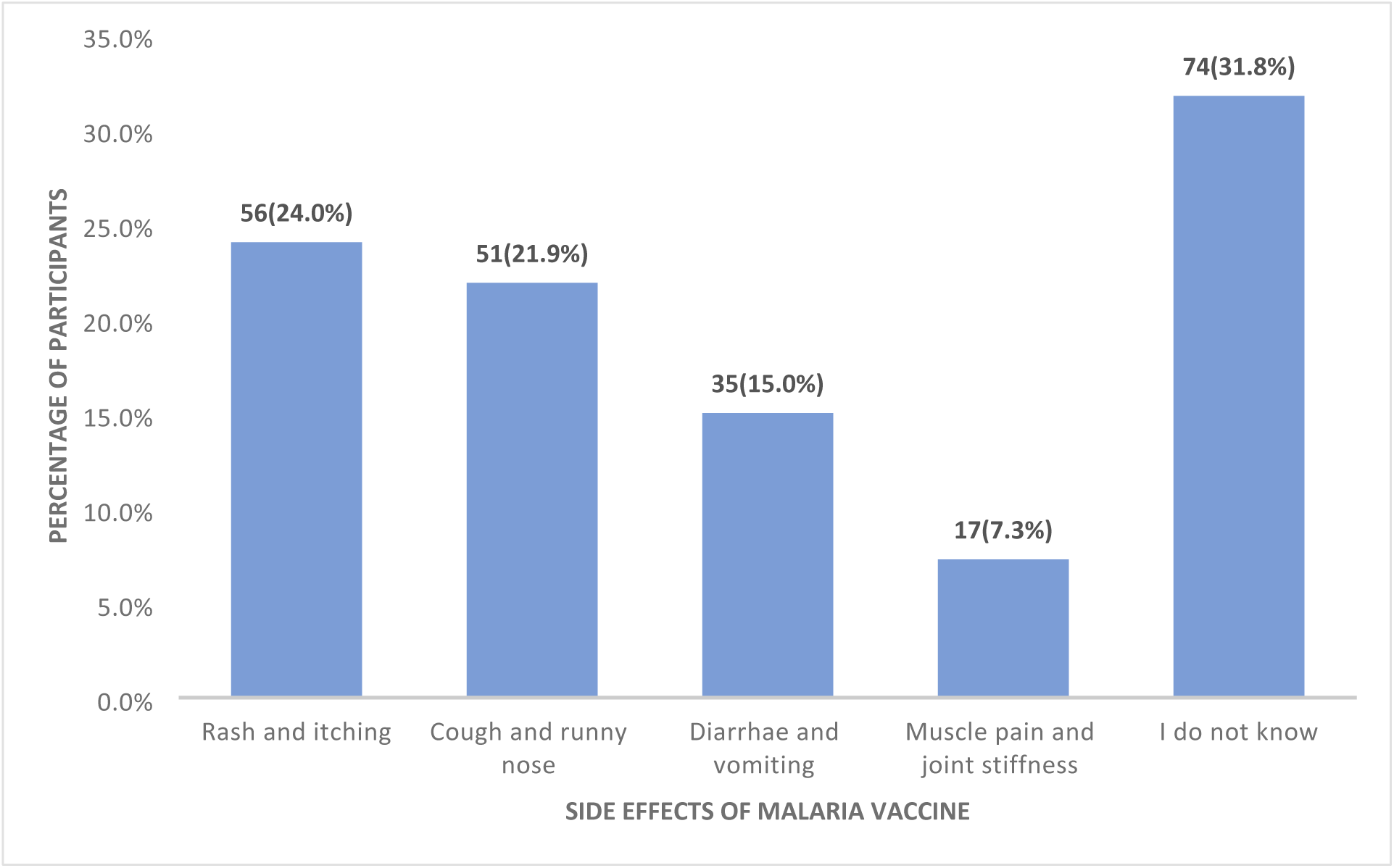
Distribution of caregivers according to their report of the side effects of malaria.

Regarding the overall knowledge of caregivers on the malaria vaccine (Figure 2), 60% of the maximum obtainable score was used as the cut off point for good knowledge. A total of seven questions were asked with a maximum obtainable score of 7. Following the 60% cutoff point, those who scored from 4 and above were classified as having overall good knowledge on the malaria vaccine and those who scored below 4 as having overall poor knowledge.

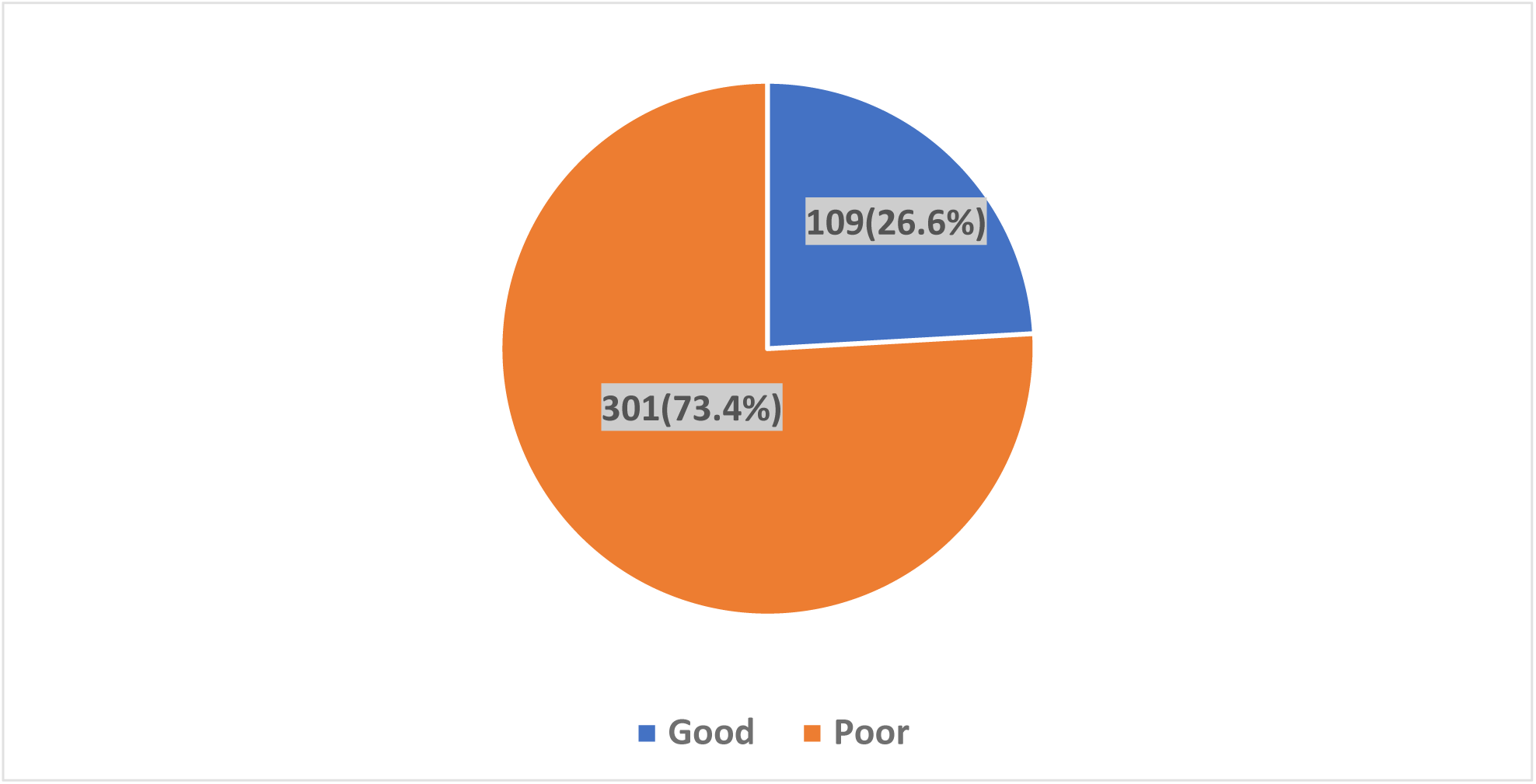

### Attitudes of Under-Five Caregivers toward the Malaria Vaccine in the Tiko Health District

Regarding the attitudes of caregivers toward the malaria vaccine (Table 3), 249 (60.7%) agreed that, vaccinating their children against malaria was crucial and 235 (57.3%) agreed that malaria vaccine was a safe option for protecting their children from malaria. A total of 219 (53.4) agreed that they trust the information provided by healthcare workers on the malaria vaccine and 169 (41.2%) agreed that completing the full course of the vaccine was essential for ensuring its effectiveness.

**Table 3:** Attitudes of under-five caregivers toward the malaria vaccine

| Variable | Strongly disagree | Disagree | Agree | Strongly agree |
| --- | --- | --- | --- | --- |
| I believe that vaccinating my child against malaria is crucial for their health | 21(5.1) | 68(16.6) | 249(60.7) | 72(17.6) |
| Malaria vaccine is a safe option for protecting my child from malaria. | 100(24.4) | 54(13.2) | 235(57.3) | 21(5.1) |
| Trust the information provided by healthcare professionals. | 24(5.9) | 137(33.4) | 219(53.4) | 30(7.3) |
| Completing the full course of malaria vaccination is essential for ensuring its effectiveness. | 130(31.7) | 67(16.3) | 169(41.2) | 44(10.7) |
| Believe that the benefits of the malaria vaccine outweigh any potential risks | 134(32.7) | 78(19.0) | 166(40.5) | 32(7.8) |
| My child should receive the malaria vaccine according to the recommended vaccination schedule. | 126(30.7) | 81(19.8) | 161(39.3) | 42(10.2) |
| I believe that the malaria vaccine is an important tool in preventing malaria related illnesses in children. | 33(8.0) | 174(42.4) | 174(42.4) | 29(7.1) |

### Overall Attitudes of Caregivers of Under Five Children Toward the Malaria Vaccine

For the overall attitudes of caregivers toward the malaria vaccine (Figure 3), a total of seven questions were asked with the options ranging from strongly disagree (0) to strongly agree (3). The maximum obtainable score was 21. A 60% cutoff point was used for overall positive attitudes. Those who scored 13 out of the 21 maximum obtainable score were classified as having overall positive attitudes and those who scored below 13 as having overall negative attitudes. Following this, 103 (25.1%) had overall positive attitudes toward the malaria vaccine.

**Figure 3:**
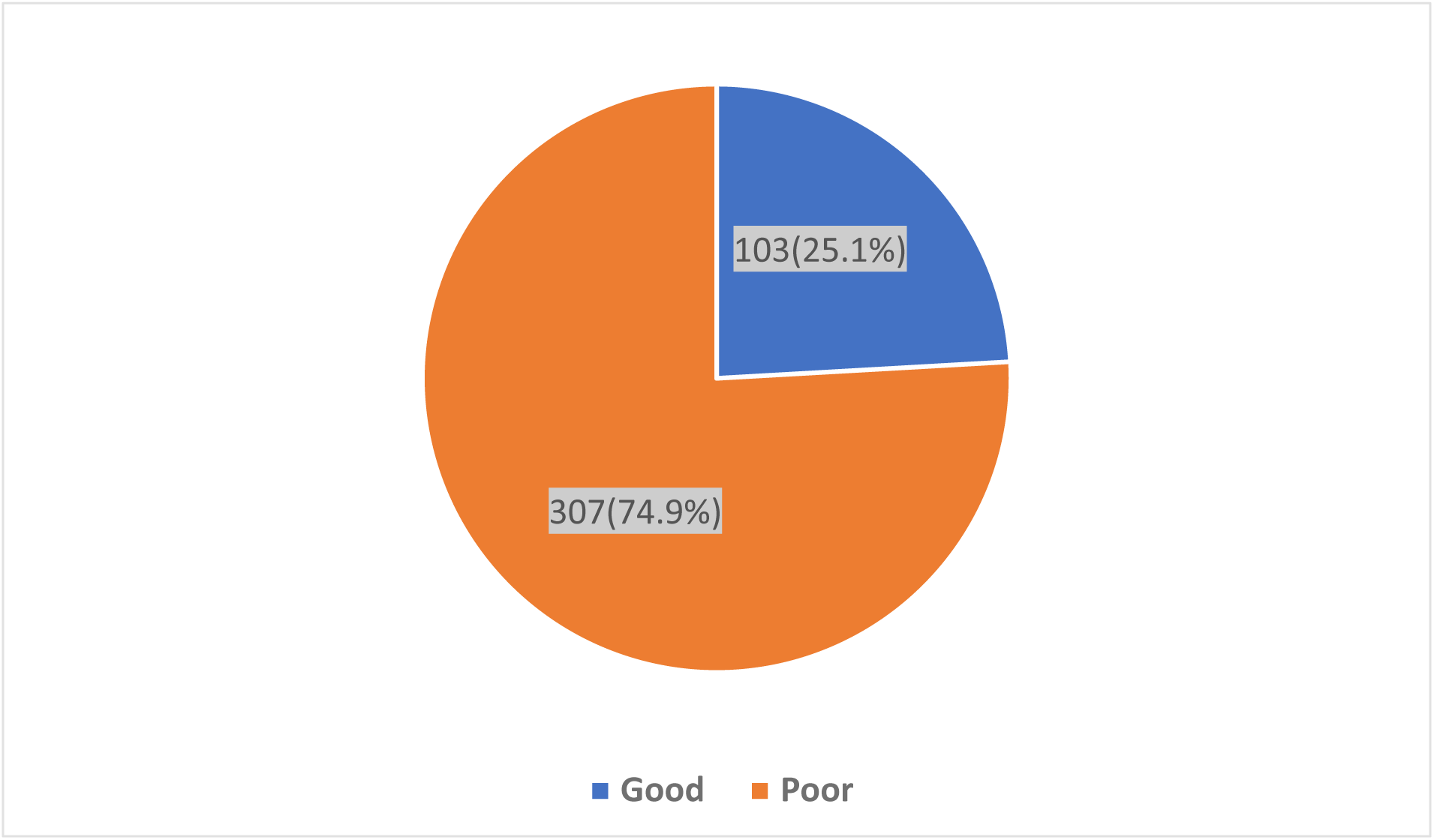
Overall attitudes of caregivers toward the malaria vaccine in the Tiko Health District.

## DISCUSSION

This study aimed to evaluate the knowledge and attitudes of caregivers of children under five years old regarding the malaria vaccine in the Tiko Health Districts. The findings reveal concerning levels of knowledge and attitudes among caregivers, which are crucial for the successful implementation of malaria vaccination programs.

### Knowledge Levels of care givers of under-five on the malaria vaccine

Only 26.6% of caregivers demonstrated good overall knowledge about the malaria vaccine. Although 60.7% of respondents had heard about the vaccine, deeper understanding was limited. A significant proportion of those aware of the vaccine did not know it had been approved in Cameroon, and many had misconceptions about its purpose—some believing it treats malaria symptoms rather than prevents infection. This low knowledge level is concerning as studies have shown that, misconception as a result of inadequate knowledge is negatively associated with the malaria vaccine uptake[21,22]. This is consistent with previous studies conducted in various regions of sub-Saharan Africa, which often report low levels of knowledge among caregivers regarding malaria vaccination. For instance, a study conducted in Tanzania in 2025 reported 14.7% awareness of caregivers on the malaria vaccine[21]. Another study in Nigeria also reported a 30% awareness of the malaria vaccine reported in Northern Nigeria[23]. However, some studies have reported slightly higher knowledge levels. For example, a study in Kenya found that around 40% of caregivers had sufficient knowledge about malaria vaccines (Muriuki et al., 2021). The disparity in knowledge levels could be attributed to differences in healthcare infrastructure, educational outreach programs, and cultural perceptions surrounding vaccination in different regions.

### Attitudes of under-five caregivers toward the malaria vaccine

Attitudinal assessment revealed that only 25.1% of caregivers had a positive attitude toward the malaria vaccine. Although a majority (60.7%) agreed that vaccinating children is crucial, concerns about safety, efficacy, and mistrust of health information remain high. Less than half of caregivers agreed that completing the full vaccine course is essential, and over 30% expressed skepticism about the vaccine’s safety.

These findings mirror results from Tanzania, where caregivers expressed concerns about vaccine side effects and effectiveness, which led to vaccine hesitancy [24]. Similarly, in Uganda, positive attitudes were reported only when caregivers had received direct counseling and community engagement interventions [25]. The relatively low trust in information from health professionals in our study (53.4% agreeing or strongly agreeing) further highlights the need for improved communication strategies that foster trust and transparency.

The low proportion of caregivers who exhibited both good knowledge and positive attitudes is particularly troubling. It suggests that many caregivers, even if aware of the vaccine, may remain hesitant due to unresolved doubts or misinformation. This disconnects between awareness and confidence can undermine vaccine rollout efforts and delay progress in malaria control.

### Implications for Public Health

These results highlight the need for intensified community sensitization efforts and caregiver-focused health education campaigns. Interventions should not only provide factual information about the vaccine but also address cultural beliefs, fears, and misinformation.

Training healthcare workers in interpersonal communication and community engagement is also essential. Given that health personnel are the most trusted sources of information, enhancing their capacity to deliver clear, empathetic, and persuasive vaccine messages could significantly influence caregiver perceptions and behaviors.

## Conclusion

In conclusion, this study reveals low levels of both knowledge and positive attitudes toward the malaria vaccine among caregivers of under-five children in the Tiko Health District. These gaps pose a significant barrier to successful vaccine implementation and require urgent, targeted interventions. Strengthening community health education, improving trust in health systems, and engaging caregivers directly will be crucial in enhancing the acceptance and uptake of the malaria vaccine in this high-risk population.

## Authors Contribution

**Conceptualization**: Idang Maureen Abiache, Divine Nsobinenyui, Chrisantus Eweh Ukah, Yunika Larissa Kumenyuy, Ngu Claudia Ngeha, Randolf Wefuan, Syveline Zuh Dang, Ndip Esther Ndip, Mirabelle Pandong Feguem, Dickson S. Nsagha

**Data Curation**: Idang Maureen Abiache, Divine Nsobinenyui, Chrisantus Eweh Ukah, Yunika Larissa Kumenyuy, Ngu Claudia Ngeha, Randolf Wefuan, Syveline Zuh Dang, Ndip Esther Ndip, Mirabelle Pandong Feguem, Dickson S. Nsagha

**Formal Analysis:** Idang Maureen Abiache, Divine Nsobinenyui, Chrisantus Eweh Ukah, Yunika Larissa Kumenyuy, Ngu Claudia Ngeha, Randolf Wefuan

**Investigation**: Chrisantus Eweh Ukah, Nicholas Tendongfor, Alan Hubbard, Elvis A. Tanue, Rasheedat Oke, Nahyeni Bassah, Sandra I. McCoy, Larissa Kumenyuy Yunika, Claudia Ngeha Ngu S. Ariane Christie, Dickson S. Nsagha, Alain Chichom-Mefire, Catherine Juillard

**Methodology:** Idang Maureen Abiache, Divine Nsobinenyui, Chrisantus Eweh Ukah, Yunika Larissa Kumenyuy, Ngu Claudia Ngeha, Randolf Wefuan, Syveline Zuh Dang, Ndip Esther Ndip, Mirabelle Pandong Feguem, Dickson S. Nsagha

**Supervision:** Abiache, Divine Nsobinenyui, Chrisantus Eweh Ukah,

**Validation:** Divine Nsobinenyui, Chrisantus Eweh Ukah, Dickson Shey Nsagha

**Visualization**: Chrisantus Eweh Ukah, Yunika Larissa Kumenyuy, Ngu Claudia Ngeha, Randolf Wefuan, Syveline Zuh Dang, Ndip Esther Ndip

**Writing – original draft:** Idang Maureen Abiache, Divine Nsobinenyui, Chrisantus Eweh Ukah, Yunika Larissa Kumenyuy, Ngu Claudia Ngeha, Randolf Wefuan, Syveline Zuh Dang, Ndip Esther Ndip, Mirabelle Pandong Feguem

**Writing – Review and Editing:** Idang Maureen Abiache, Divine Nsobinenyui, Chrisantus Eweh Ukah, Yunika Larissa Kumenyuy, Ngu Claudia Ngeha, Randolf Wefuan, Syveline Zuh Dang, Ndip Esther Ndip, Mirabelle Pandong Feguem, Dickson S. Nsagha

## Data Availability

The dataset supporting the conclusions of this article is available from the corresponding author upon reasonable request. Due to ethical restrictions related to participant confidentiality and the conditions imposed by the ethical review board that approved the study (Southwest Regional Ethics Committee, Cameroon), the data cannot be made publicly available. De-identified data may be shared upon request and with appropriate institutional approvals.

